# Clinical characteristics and epidemiology survey of lung transplantation recipients accepting surgeries during the COVID-19 pandemic : from area near Hubei Province

**DOI:** 10.1101/2020.07.06.20147264

**Authors:** Lingxiao Qiu, Shan shan Chen, Cong Wang, Caihong Liu, Huaqi Wang, Xiaoguang Zhao, Zeming Fang, Siyuan Chang, Gaofeng Zhao, Guojun Zhang

## Abstract

Lung transplantation recipients (LTx) were susceptible to severe acute respiratory syndrome-corona virus-2 (SARS-Cov-2) and suffered a higher mortality risk than healthy subjects. Here we aim to analyze whether it was appropriate or and valuable to maintain lung transplant programs in medical institutions accepting coronavirus disease 2019 (COVID-19) patients. In this study, the clinical characteristics, laboratory testing and epidemiology survey results of 10 LTx recipients undergoing allograft lung transplantation surgeries in the First Affiliated Hospital of Zhengzhou University during the COVID-19 pandemic were collected. A web-based epidemiology questionnaire was used to collect the information of LTx recipients after discharge. Up to now, none of the LTx recipients or their family members get infected with SARS-CoV-2 during the novel coronavirus pandemic. In conclusion, under the premise of taking appropriate preventive measures during hospitalization and after discharge, the lung transplant program can be maintained in the medical institution that accepts patients with COVID-19.

## 1. Introduction

Since the outbreak of severe acute respiratory syndrome coronavirus 2 (SARS-Cov-2) in Wuhan, Hubei, China in December 2019, large amounts of people have been infected and killed due to the rapid spread of such virus in China and around the world^[1, 2]^. SARS-CoV-2 was previously called 2019-nCoV, until February 11, 2020, the International Committee on Taxonomy of Viruses renamed this virus^[3]^. At the same time, World Health Organization(WHO)renamed the disease caused by SARS-CoV-2 as Coronavirus Disease 2019 (COVID-19). SARS-CoV-2 targets angiotensin-converting enzyme 2 (ACE2) as a receptor and enters the host cell through it, resulting in a novel CoV-related pneumonia ^[4]^. According to real-time statistics released by Johns Hopkins University, until May 5th, 2020, there were more than 3.87 million confirmed cases of COVID-19 globally, and the death toll exceeded 250 thousand, which reached to 250687 cases.

Organ transplantation recipients are a special group of population with immunity greatly suppressed due to the immunosuppressants. Compared to healthy individuals, they are more susceptible to SARS-CoV-2 and at even higher risk of severe illness from COVID-19 according to “list of susceptible people” published by Public Health England ^[5]^. It is worth noting that current epidemiological evidence indicates that most of the patients with COVID-19 are not severe cases and enable to recover after appropriate treatment. However, the mortality rate of COVID-19 patients with comorbidities (especially with lung diseases) was much higher than those without diseases ^[6, 7]^.

During the pandemic of COVID-19, large amounts of transplantation surgeries were delayed or canceled, especially in the seriously affected areas. Our medical institution is located in Henan Province in the central of China and adjacent to Wuhan, Hubei Province. During the COVID-19 pandemic, our area was severely affected. At this critical time, whether the lung transplant program needs to be maintained, and the clinical characteristics and SARS-CoV-2 infection conditions of these lung transplantation (LTx) recipients remain unclear. In this work, we collected characteristics of these LTx recipients and designed an epidemiology questionnaire to obtain the information of SARS-CoV-2 infection condition and after-discharge preventive measures of LTx recipients who underwent surgeries in the hospital accepting COVID-19 patients during the COVID-19 pandemic.

## 2. Materials and methods

### 2.1 Data source

This is a retrospective and single-center study, all data sources coming from the First Affiliated Hospital of Zhengzhou University of China.

### 2.2 Selection criteria

All patients receiving allograft lung transplantation surgeries in the First Affiliated Hospital of Zhengzhou University were included, from the inception of the COVID-19 pandemic (January 2020) to the day when the Secondary Prevention Level of Public Health Emergencies in Henan Province was removed (May 5^th^, 2020).

### 2.3 Statistical analyses

A web-based epidemiology questionnaire was used to collect the information of LTx recipients after discharge, which was designed by clinicians and public health experts and collected by 2 independent investigators.

The continuous variable of a normal distribution is expressed as mean value (standard deviation), and the continuous variable that is not normally distributed is expressed as median (interquartile space). Categorical variables were described through count and frequency. All statistical analyses were conducted on version 26.0 of SPSS software.

## 3. Results

### 3.1 Patients’ characteristics

A total of 10 LTx recipients consisting of 1 female and 9 males were identified in this study, whose characteristics were showed in **Table 1**. The median age is 60 (50∼64) and the youngest case is only 15 years old. The main cause of lung transplantation in this study was idiopathic interstitial pneumonia (IIP) accounting for 60% and one rare cause was cystic fibrosis with only one case. All patients suffered dyspnea on admission. From the time of admission to accept lung transplantation surgery, 6 patients received ordinary nasal catheter oxygen therapy, 1 patient received high-flow nasal catheter oxygen therapy (HFNC) with 60L/min, and 3 patients underwent endotracheal intubation combined with an invasive ventilator for assisted breathing. In general, patients with lung fibrosis suffered severer dyspnea than those who suffered bronchiectasis. As for surgical approaches, single lung transplantation surgery was the main surgical method used in this study. Only one 15-year patient received bilateral lung transplantation surgery, who was diagnosed as cystic fibrosis. Meanwhile, part of the LTx recipients suffered comorbidities, including 1 case with viral hepatitis of type B and 1 case with diabetes, hyperlipidemia and subclinical hyperthyroidism.

**Table 1.** Characteristics of included recipients.

| **Variables** | **Recipients (n=10)** |
| --- | --- |
| **Age** |  |
| ≤35 | 1 (10%) |
| 36-60 | 4 (40%) |
| 61-70 | 5 (50%) |
| **Gender** |  |
| Male | 9 (90%) |
| Female | 1 (10%) |
| **Marriage** |  |
| Married | 9 (90%) |
| Unmarried | 1 (10%) |
| Divorced | 0 |
| Widowed | 0 |
| **Reasons for lung transplantation** |  |
| IIP | 6 (60%) |
| Bronchiectasis | 2 (20%) |
| Others | 2 (20%) |
| **Surgical approach** |  |
| Right single lung transplantation | 4 (40%) |
| Left single lung transplantation | 5 (50%) |
| Bilateral lung transplantation | 1 (10%) |
| **respiratory assistant ventilation** |  |
| Ordinary nasal catheter | 6 (60%) |
| HFNC | 1 (10%) |
| Invasive ventilator | 3 (30%) |
| **Comorbidity** |  |
| Hyperlipidemia | 1 (10%) |
| Diabetes | 1 (10%) |
| Subclinical hyperthyroidism | 1 (10%) |
| viral hepatitis type B | 1 (10%) |

Median age (IQR) 60 years (50~64)
IQR= Interquartile range
ECMO= Extracorporeal membrane oxygenation
IIP= Idiopathic interstitial pneumonia
HFNC= High-flow nasal cannula oxygen therapy
COPD= Chronic obstructive pulmonary disease
Data are expressed as n/N ( % )

### 3.2 Laboratory results

Laboratory results of the 10 LTx recipients before lung transplantation surgery were summarized in **Table 2**, including blood routine, biochemistry, myocardial enzyme, blood gas analysis, and tacrolimus metabolic type. In all identified patients, the average white blood cell (WBC) count was 9.5±3.9×109 cells/liter and 3 (30%) cases had an abnormal WBC count. The average neutrophil percentage was 70.4%±15.2% with 3 cases abnormal. The average lymphocyte percentage was 19.2%±11.2% and 4 (40%) LTx recipients had lymphopenia, among them 2 cases had a lymphocyte percentage lower than 10%. Besides, 1 patient had both liver and kidney function insufficiency and 3 patients suffered heart function insufficiency before accepting lung transplantation surgery and taking tacrolimus. Blood gas analysis results indicated all the 10 patients appeared respiratory failure, including 2 cases of type □ respiratory failure and 8 cases of type □ respiratory failure. Due to the personal financial situation and medical insurance, tacrolimus metabolic type tests were only conducted on 5 LTx recipients. The results showed that 1 subject had a normal metabolism and 4 subjects had a slow metabolism.

**Table 2.** Laboratory results of recipients before lung transplantation surgery.

| **Laboratory results** | **Recipients**<br>**(n=10)** |
| --- | --- |
| **Blood routine** |  |
| WBC count ($\times 10^9$ cells/liter) | 9.5±3.9 |
| Abnormal WBC count | 3(30%) |
| Neutrophil count ($\times 10^9$ cells/liter) | 7.0±4.2 |
| Abnormal neutrophil count | 3(30%) |
| Lymphocyte count ($\times 10^9$ cells/liter) | 1.7±1.1 |
| Abnormal lymphocyte count | 5(50%) |
| Lymphopenia count | 4(40%) |
| Lymphocytosis count | 1(10%) |
| Neu% | 70.4±15.2 |
| Abnormal Neu% count | 3(30%) |
| Lym% | 19.2±11.2 |
| Abnormal Lym% count | 6(60%) |
| <10% | 2(20%) |
| 10%~19% | 4(40%) |
| 20%~50% | 4(40%) |
| **Biochemistry** |  |
| ALT (U/liter) | 23.6±29.6 |
| AST (U/liter) | 25.4±21.5 |
| Urea (mmol/liter) | 5.9±2.9 |
| Cr (mmol/liter) | 49.2±10.3 |
| **Myocardial enzyme** |  |
| NT-proBNP (pg/mL) | 728.7±1332.8 |
| Abnormal NT-proBNP count | 3(30%) |
| <300 | 7(70%) |
| 300~2999 | 2(20%) |
| 3000~5000 | 1(10%) |
| **Blood gas analysis** |  |
| PH | 7.4±0.1 |
| PaO<sub>2</sub> (mmHg) | 74.0±14.4 |
| PaCO<sub>2</sub> (mmHg) | 56.5±16.2 |
| HCO<sub>3</sub><sup>-</sup> ( mmol/L ) | 32.1±7.8 |
| Types□respiratory failure count | 2(20%) |
| Types□ respiratory failure count | 8(80%) |
| Respiratory acidosis count | 6(60%) |
| Respiratory acidosis combined<br>with metabolic alkalosis count | 2(20%) |
| **Tacrolimus metabolic types** |  |
| Fast metabolism | 0 |
| Normal metabolism | 1(10%) |
| Slow metabolism | 4(40%) |
| Unknown | 5(50%) |

WBC=White blood cell
Neu= Neutrophil
Lym= Lymphocyte
ALT= Alanine aminotransferase
AST= Aspartate transaminase
Cr= Creatinine
NT-proBNP= N terminal pro B type natriuretic peptide
PaO<sub>2</sub>= partial pressure of oxygen in artery
PaCO<sub>2</sub>= partial pressure of carbon dioxide in artery

### 3.3 Epidemiologic survey

A total of 6 LTx recipients were followed up and all of them lived in Henan Province after discharge. The epidemiological survey results of SARS-CoV-2 prevention in these LTx recipients after discharge were presented in **Table 3**. Good hand hygiene habitats were kept in all followed-up LTx recipients, and all of the LTx recipients (100%) consciously often or occasionally used the seven-step hand-washing method to wash hands. It should be noted that all included LTx recipients owned personal hand towels. Indoor ventilation measures were daily carried out in all LTx recipients’ homes. Masks were essential especially the medical/surgical masks (83%) if these LTx recipients went out. 50% of the LTx recipients tended to choose two types of masks, while the other 50% wore only one type of mask. Indoor disinfection behaviors were conducted in 5 LTx recipients’ homes (83%), including floors, the surface of furniture and toilets. Due to the demand for postoperative recovery and the prevention of pathogenic bacteria and viruses, separate bedroom (100%), personal bedsheets/ quilts (100%) and drinking glasses (100%) were prepared for all these LTx recipients. At the end of follow-up, none of these LTx recipients and their family members were infected with SARS-CoV-2 during the coronavirus pandemic.

**Table 3.** Epidemiology survey on SARS-CoV-2 prevention after discharge.

| **Items** | **Recipients**<br>**(n=6)** |
| --- | --- |
| **Hand hygiene** |  |
| Returning home | 5(83%) |
| Before eating food | 5(83%) |
| After toilets | 6(100%) |
| After coughing or sneezing | 4(67%) |
| **Hand hygiene approaches** |  |
| Seven-step hand-washing method | 6(100%) |
| Often | 2(33%) |
| Occasionally | 4(67%) |
| **Hand hygiene products** |  |
| Soaps | 4(67%) |
| Hand sanitizer | 3(50%) |
| Dry hand paper | 3(50%) |
| Personal hand towel | 5(83%) |
| **Open windows for ventilation** |  |
| Frequency/Daily |  |
| 1-3 | 5(83%) |
| > 3 | 1(17%) |
| Time/ Each |  |
| ≤30 minutes | 3(50%) |
| > 30 minutes | 3(50%) |
| **Masks** |  |
| Ordinary masks | 2(33%) |
| Medical/ Surgical masks | 5(83%) |
| N95 masks | 2(33%) |
| **Indoor disinfection** |  |
| Floor | 4(67%) |
| Furniture surface | 5(83%) |
| Toilets | 4(67%) |
| **Living situation** |  |
| Separate bedroom | 6(100%) |
| Separate toilet | 1(17%) |
| Shared toilet | 5(83%) |
| **Household items** |  |
| Personal drinking glass | 6(100%) |
| Personal tableware | 4(67%) |
| Personal bedsheets/quilts | 6(100%) |

Data are expressed as n (%) and n/N (%).

## 4. Discussion

This is the largest report of LTx patients accepting allograft lung transplantation surgery in severe epidemic areas during the COVID-19 pandemic. In our study, part of LTx recipients had hyperlipidemia, diabetes, subclinical hyperthyroidism, viral hepatitis type B comorbidities. Besides, 4 LTx recipients (40%) had lymphopenia, 3 LTx recipients (30%) had impaired heart function and 1 LTx recipient had both of the impaired alanine aminotransferase (ALT) and aspartate transaminase (AST).

Previous studies have indicated that human beings are susceptible to SARS-Cov-2^[8, 9]^. The immunity of LTx recipients is greatly suppressed because of immunosuppressive agents, making them more susceptible to SARS-COV-2 infection^[5]^. Early COVID-19 patients’ reports indicated that lymphopenia and impaired ALT were associated with severe complicated disease and even resulted in death^[10]^. Recent SARS-CoV-2 patients’ reports also showed that age ≥ 65 years, dyspnea, coronary heart disease, cerebrovascular disease and AST level > 40 U/L were independent risk factors associated with fatal outcome of infective subjects^[11]^. All included subjects in our study were susceptible people of SARS-CoV-2, and they would suffer a higher mortality rate if they were infected.

By the end of the follow-up, no LTx recipients included in our reports have been infected by SARS-CoV-2. We surveyed SARS-CoV-2 preventive measures taken by these LTx recipients after discharge, based on a web comprehensive questionnaire. Results revealed that the seven-step hand-washing method was done (100%) by all the LTx recipients and masks were protective items that must be carried when they went out (100%). Separate bedrooms (100%), personal bedsheets/ quilts (100%) and drinking glass (100%) were also prepared for them by their family members. The rate of indoor disinfection behavior reached 83%. Consistent with previous reports^[12, 13]^, the key reasons for the zero SARS-CoV-2 infective rate of LTx recipients in our study may be due to the good performance of hand hygiene, wearing masks and indoor disinfection, and even isolation from family members. According to the guideline published by the World Health Organization (WHO), using masks can reduce potential exposure risk from an infected patient during the pre-symptomatic or an asymptomatic phase^[14]^. Some clinical trials have shown that masks and hand hygiene could effectively protect the coronavirus transmission in the community^[15]^. Besides, masks seemed to be more effective than hand hygiene alone, and the two together are more protective^[16]^.

In our reports, 83% of the LTx recipients tended to use medical masks/ surgical masks or N95 masks rather than ordinary masks. However, the WHO stated that medical masks were necessary only when people were involved in taking care of people with suspected or confirmed COVID-19^[17]^. Since SARS-CoV-2 outbroke in December 2019, it had caused considerable psychological stress on both public and doctors and led to the increasing mental health problems^[18-21]^. The abovementioned behavior precisely reflected the panic of the epidemic among the public. Interestingly, whether it was necessary to wear higher-level protective masks for people who received lung transplantation surgery during the COVID-19 pandemic was unknown. However, the use of medical masks, good hand hygiene behaviors and frequent disinfection of indoor furniture surfaces can help to release the LTx recipients’ anxieties during this severe period.

It should be noted that anxieties in part of LTx recipients were observed during our follow-up, which mostly came from the exposure to multiple co-occurring risk factors (e.g., spouse’s or children’s job loss, credit-card repayment, marital conflict). It can be seen that, in addition to the impact of the disease on public health, this large-scale public health crisis has caused incalculable damage to the global and national economy because of the nationwide shutdown and home confinement directives that prohibit national and regional economic activities^[22, 23]^. We then separate this investigation in the future.

Since Henan Province is close to Hubei Province, therefore, our area was severely affected during the COVID-19 pandemic. Our hospital was the main medical institution to treat COVID-19 patients in Henan Province. To ensure the maintenance of the transplant programs, patients were admitted to the hospital only when no imaging findings of SARS-CoV-2 infection on chest CT or tested negative for SARS-CoV-2. Besides, our medical institution set an independent ward for SARS-CoV-2 patients and a specific ICU for transplant patients. These two wards are not in the same building. SARS-CoV-2 prevention team was also established to be responsible for hospital-acquired infection. All LTx recipients and their family members were educated about COVID-19 during hospitalization, including the source of infection, route of transmission, susceptible population and personal protection procedures. All these measures played positive roles in protecting these LTx recipients from SARS-CoV-2 infection.

There are several limitations to the current study. Firstly, this is a single-center study. Secondly, the SARS-CoV-2 is not cleared in China and other countries of the world, these LTx recipients still face the risk of infection. Therefore, observation and follow-up are warranted. Furthermore, a larger sample size will help to improve the greater reliability of the data.

In conclusion, according to our survey, if the medical institution has sufficient wards, medical staff, appropriate measures of preventing nosocomial infections and also can provide LTx recipients with correct education on the prevention of SARS-CoV-2 infection, we believe that it is feasible to maintain lung transplant program in severely affected areas during the COVID-19 pandemic.

## Data Availability

The data that support the findings of this study are available from the corresponding author upon reasonable request.

## References

1. Li, Q., et al., Early Transmission Dynamics in Wuhan, China, of Novel Coronavirus-Infected Pneumonia. N Engl J Med, 2020. 382(13): p. 1199–1207.

2. Huang, C., et al., Clinical features of patients infected with 2019 novel coronavirus in Wuhan, China. Lancet, 2020. 395(10223): p. 497–506.

3. Viruses, C.S.G.o.t.I.C.o.T.o., The species Severe acute respiratory syndrome-related coronavirus: classifying 2019-nCoV and naming it SARS-CoV-2. Nat Microbiol, 2020. 5(4): p. 536–544.

4. Zhou, P., et al., A pneumonia outbreak associated with a new coronavirus of probable bat origin. Nature, 2020. 579(7798): p. 270–273.

5. England, P.H., Guidance on social distancing for everyone in the UK, 30 March 2020.

6. Guan, W.J., et al., Comorbidity and its impact on 1590 patients with COVID-19 in China: a nationwide analysis. Eur Respir J, 2020. 55(5).

7. Liang, W.H., et al., Clinical characteristics and outcomes of hospitalised patients with COVID-19 treated in Hubei (epicentre) and outside Hubei (non-epicentre): a nationwide analysis of China. Eur Respir J, 2020. 55(6).

8. Gussow, A.B., et al., Genomic determinants of pathogenicity in SARS-CoV-2 and other human coronaviruses. Proc Natl Acad Sci U S A, 2020.

9. Shang, J., et al., Cell entry mechanisms of SARS-CoV-2. Proc Natl Acad Sci U S A, 2020. 117(21): p. 11727–11734.

10. Peiris, J.S., et al., Coronavirus as a possible cause of severe acute respiratory syndrome. Lancet, 2003. 361(9366): p. 1319–25.

11. Chen, R., et al., Risk Factors of Fatal Outcome in Hospitalized Subjects With Coronavirus Disease 2019 From a Nationwide Analysis in China. Chest, 2020.

12. Brauer, M., et al., Global Access to Handwashing: Implications for COVID-19 Control in Low-Income Countries. Environ Health Perspect, 2020. 128(5): p. 57005.

13. Gharpure, R., et al., Knowledge and Practices Regarding Safe Household Cleaning and Disinfection for COVID-19 Prevention - United States, May 2020. MMWR Morb Mortal Wkly Rep, 2020. 69(23): p. 705–709.

14. World Health, O., Advice on the use of masks in the context of COVID-19: interim guidance, 5 June 2020.

15. Leung, N.H.L., et al., Respiratory virus shedding in exhaled breath and efficacy of face masks. Nat Med, 2020. 26(5): p. 676–680.

16. MacIntyre, C.R. and A.A. Chughtai, A rapid systematic review of the efficacy of face masks and respirators against coronaviruses and other respiratory transmissible viruses for the community, healthcare workers and sick patients. Int J Nurs Stud, 2020. 108: p. 103629.

17. World Health, O., Rational use of personal protective equipment for coronavirus disease (COVID-19) and considerations during severe shortages: interim guidance, 6 April 2020.

18. Jungmann, S.M. and M. Witthöft, Health anxiety, cyberchondria, and coping in the current COVID-19 pandemic: Which factors are related to coronavirus anxiety? J Anxiety Disord, 2020. 73: p. 102239.

19. Wade, M., H. Prime, and D.T. Browne, Why we need longitudinal mental health research with children and youth during (and after) the COVID-19 pandemic. Psychiatry Res, 2020. 290: p. 113143.

20. Wang, C., et al., Immediate Psychological Responses and Associated Factors during the Initial Stage of the 2019 Coronavirus Disease (COVID-19) Epidemic among the General Population in China. Int J Environ Res Public Health, 2020. 17(5).

21. Rosenbaum, L., Facing Covid-19 in Italy - Ethics, Logistics, and Therapeutics on the Epidemic’s Front Line. N Engl J Med, 2020. 382(20): p. 1873–1875.

22. Ceylan, R.F., B. Ozkan, and E. Mulazimogullari, Historical evidence for economic effects of COVID-19. Eur J Health Econ, 2020: p. 1–7.

23. Nicola, M., et al., The socio-economic implications of the coronavirus pandemic (COVID-19): A review. Int J Surg, 2020. 78: p. 185–193.

